# Phylodynamic inference of the contribution of transmission routes in infectious disease outbreaks

**DOI:** 10.1101/2025.06.17.25329759

**Authors:** Bastiaan R. Van der Roest, Don Klinkenberg, Egil A.J. Fischer, Martin C.J. Bootsma, Mirjam E.E. Kretzschmar

## Abstract

Understanding which types of contact drive pathogen transmission is critical for outbreak control. Traditionally, inference of transmission events with contact tracing or network analyses is used to identify high-risk contacts. Inference based on genetic data might offer higher accuracy on identifying the true events, but is rarely used to evaluate the contribution of different types of contact to transmission. We extended the Bayesian phylodynamic model `phybreak` to incorporate data on different types of contacts between cases. The model estimates the fraction of transmission events attributable to different types of contact by combining pathogen genetic sequences, sampling times, and contact data. We evaluated performance through simulations across various scenarios — different prevalences of contact types, independent versus correlated types — and applied the method to the 2020 SARS-CoV-2 outbreak in Dutch mink farms. Simulations showed that when contacts of a specific type were sparse but important for transmission, the model accurately estimated the number of transmissions attributable to that type of contact. Performance declined with more prevalent or more positively correlated contact types. Contact data improved transmission tree inference, particularly under conditions where genetic data alone lacked resolution. When applying the method to the SARS-CoV-2 outbreak, we estimated that in 76% of all transmission events between farms linked via shared personnel, transmission occurred through a link of this type. Veterinary service providers and feed suppliers were less strongly associated with transmission. This method allows for simultaneous inference of transmission trees and quantification of the importance of contact types using both genetic and structured contact data. It improves accuracy of inferring who infected whom under realistic conditions and is applicable to a range of outbreak settings. These findings can inform intervention and surveillance strategies in both human and animal populations.

**Author summary:** In outbreaks of infectious diseases, there are often many possible ways a pathogen can be transmitted — through direct contact, e.g. skin-to-skin contact, or indirect contact, e.g. shared health-care workers, service providers or environments. However, it is hard to know which of these connections actually matter most for transmission, although this can be crucial to stop the spread of the disease. In this study, we created a method that uses both genetic data from a pathogen and records of contacts between cases to investigate which connections contributed most to spreading a disease.

We tested our approach using simulated outbreaks and applied it to the spread of COVID-19 among Dutch mink farms. We found that our method worked well on simulated outbreaks, when the contacts were not too common. In the example of COVID-19 in mink farms, we found that shared personnel was the most risky type of contact, more so than shared feed suppliers or veterinary service providers. This insight can help authorities to focus their intervention efforts on the most risky contacts in future outbreaks. Although we have shown an example of an outbreak among animals, our method can also be used for human infectious disease outbreaks.

## Introduction

During an infectious disease outbreak, the pathogen of concern may be transmitted via multiple types of contact. These types range from direct, e.g. skin-to-skin, contact to indirect contact e.g via health-care workers or shared personnel in case of an animal disease. Knowledge of the contribution of the different contact types to transmission is pivotal for public health authorities to design effective interventions [1].

Transmission routes are defined as the transmission of a pathogen from an infector to its infectee via a type of contact. Conventionally, these routes are determined by epidemiological approaches using contact tracing. Risk factors are classified based on data on contacts made by infected individuals, and the infection status of their contactees [2] [3]. In the absence of contact data of uninfected individuals, types of contact could be classified as high-risk based on the contacts between infected cases only by estimating transmission probabilities for the types of contacts. These methods are effective in identifying important transmission routes in the contact of an outbreak investigation. Building on contact tracing results, social networks analysis maps relationships between individuals or groups, such as patients, wards or farms, as nodes connected by potential transmission events. This is especially useful for capturing both direct and indirect contacts, such that the true importance of all contact types is inferred [4].

In the last decades, genetic data have emerged as a powerful source for inferring transmission dynamics. Using the genetic sequences of pathogens, reconstruction of transmission trees can reveal how an infection has spread through a population. These methods provide evidence for who infected whom even when contact tracing data are not available [5] [6].

Inference of transmission trees using both genetic data and contact data seems promising to estimate the relative contributions of different transmission routes, using the specification of types of contact between cases and the higher resolution of the genetic data. Previous effort to integrate both data sources used a kernel approach, based on the pathogen genotype and epidemiological data such as spatial distance or contact [7]. Other methods used a sequential approach: first, transmission trees were inferred from genetic data; subsequently, contacts between infector-infectee pairs were analysed to identify high-risk transmission routes, such as human-mediated transport [8] or wind dispersal [9] in the context of Highly Pathogenic Avian Influenza. Furthermore, social network analyses have been used in combination with phylogenetic trees to infer transmissions, as done for an outbreak of Tuberculosis [10]. An alternative method for integrating genetic and contact data in inference was proposed by Campbell et al. [11], who extended the transmission tree inference model `outbreaker2` by modelling contact tracing as a probabilistic process. Contact data were shown to improve the inference of the transmission tree. In that way, conflicting data, such as potential transmission events supported by contact data but with high genetic diversity, are treated more systematically.

We propose to integrate contact data into transmission tree inference in such a way that the importance of different types of contact can be quantified. We aim to do this for multiple contact types in the same inference. To achieve this aim, we extended the Bayesian transmission tree inference method `phybreak` [12], such that this method can distinguish transmission events that have occurred through different types of contact. Through simulation studies, we evaluated the model’s performance in estimating the importance of one or multiple known types of contact. As a real-world application, we analysed a SARS-CoV-2 outbreak in Dutch mink farms [13] [14] to demonstrate the utility of this approach and gain insights into how different types of contact contributed to spread of SARS-CoV-2 among mink farms in the Netherlands.

## Results

We consider an outbreak of an infectious disease, for which we know all cases - infected hosts - with their detection times and pathogen sequences. There is no information about susceptible hosts. We further have contact data, indicating for all pairs of hosts, for one or more types of contact, whether they were observed. Types of contact are considered along which transmission can have taken place, so the types may differ between infectious diseases. For respiratory diseases these could be physical contact or being in close proximity, for livestock diseases this could be farm-to-farm movement of animals or sharing a veterinary service provider. We assume that all contacts of a known type are reported, that they are undirected and that they were always present during the outbreak. Hosts may have been infected by infection from outside the target population (introduction), by an infector through any of the known types of contact (known transmission route) or by an infector through an unknown type of contact (unknown transmission route) (see Figure 1). We treat the unknown route as a separate route, with all pairs of hosts connected.

**Fig 1.**
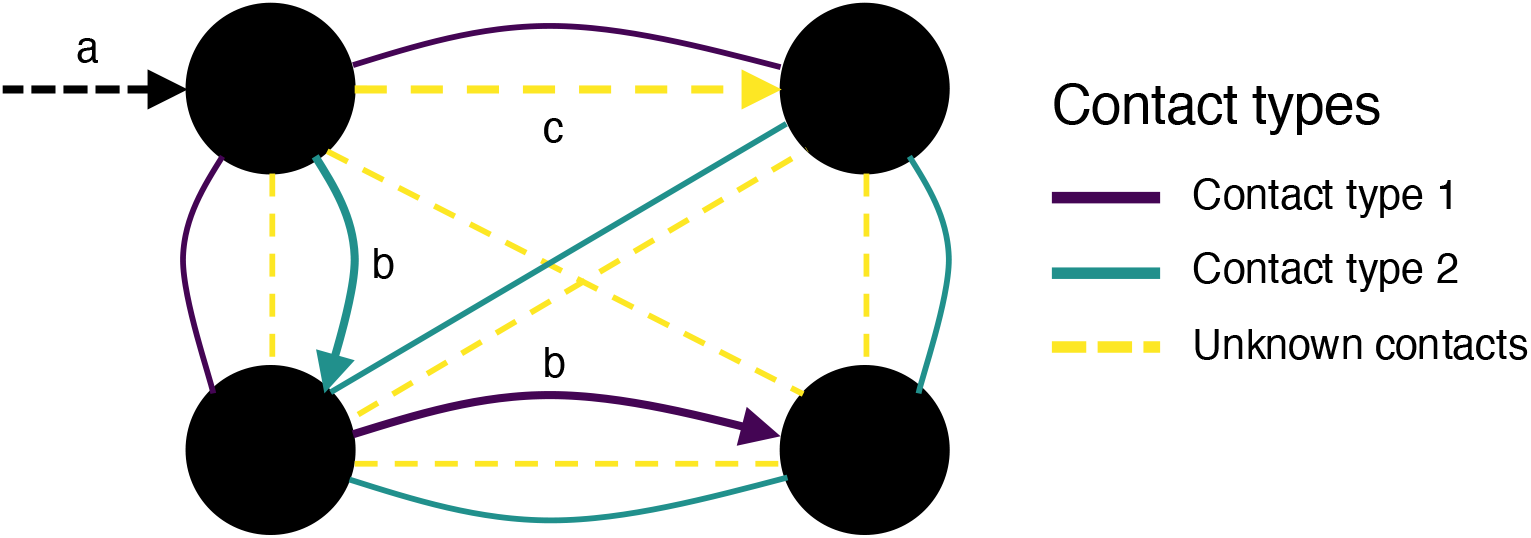
Outbreak with 3 infection routes, defined by the contact type. Arrows represent infection of the hosts (black dots). (a) Infection from outside the target population: introduction. (b) Infection through any of the known contact types (known transmission route). (c) Infection through an unknown type of contact (unknown transmission route)

### Transmission tree inference

Transmissions in the population of infected hosts are inferred using a Bayesian model, where the likelihood of observing these transmissions is informed by epidemiological and genetic links between hosts and samples [14] (see Methods). Our main result is that we provide a method that includes contact data in the `phybreak` model. To achieve this, the likelihood for transmission was extended by adding the occurrence of contact of different types between hosts and the importance of the type of contact for transmission. This importance for transmission is quantified by estimating the expected fraction of transmissions per transmission route using the type of contact.

### Simulation-based assessment of contact contribution estimates

By simulation, we constructed contact data matrices for 25 simulated outbreaks with 20 hosts, for three groups of scenarios. Each outbreak was simulated with one introduction and we let the model estimate the number of introductions. In scenario group 1 we constructed a single contact data matrix with varying importance of the known contact type, determined by the number of transmissions that have occurred via the corresponding transmission route, and the proportion of the contact type in the population (Table 1). The model estimated the number of transmissions via the known transmission route to be close to the simulated number of transmissions when the known contact is sparse, i.e., the proportion of contacts of this type in the population is low (Fig 2A). For increasing proportions of this type of contact, the number of transmissions approaches half of the total transmissions. For the case that every host had contact with every other host via a contact of this type (proportion of known contact is 1), there is no information to distinguish the known from the unknown route. So, the inferred transmissions were equally divided over the known and unknown transmission routes.

**Table 1.** Groups of simulation scenarios for contact data matrices and the used values of proportion of contact *p* and number of transmissions via the route *x*.

| Scenario's | Definition | Proportion of contact $p$ | Number of transmissions via route |
| --- | --- | --- | --- |
| Group 1 | 1 contact type |  |  |
| | random contacts | $p = 0.1, 0.5, 1$ | $x = 0, 10, 19$ |
| Group 2 | 2 contact types | $(p^1, p^2) = (0.1, 0.1)$ | $(x_1, x_2) = (19, 0), (15, 4),$ |
| | random contacts | $(0.5, 0.5), (1, 1)$ | $(5, 5)$ |
| Group 3 | 2 contact types<br>(anti-)correlated contacts | $(p^1, p^2, p^{12}) = (0.1, 0.1, 0), (0.1, 0.1, 0.1)$ | $(x_1, x_2) = (15, 4), (4, 0)$ |

**Fig 2.**
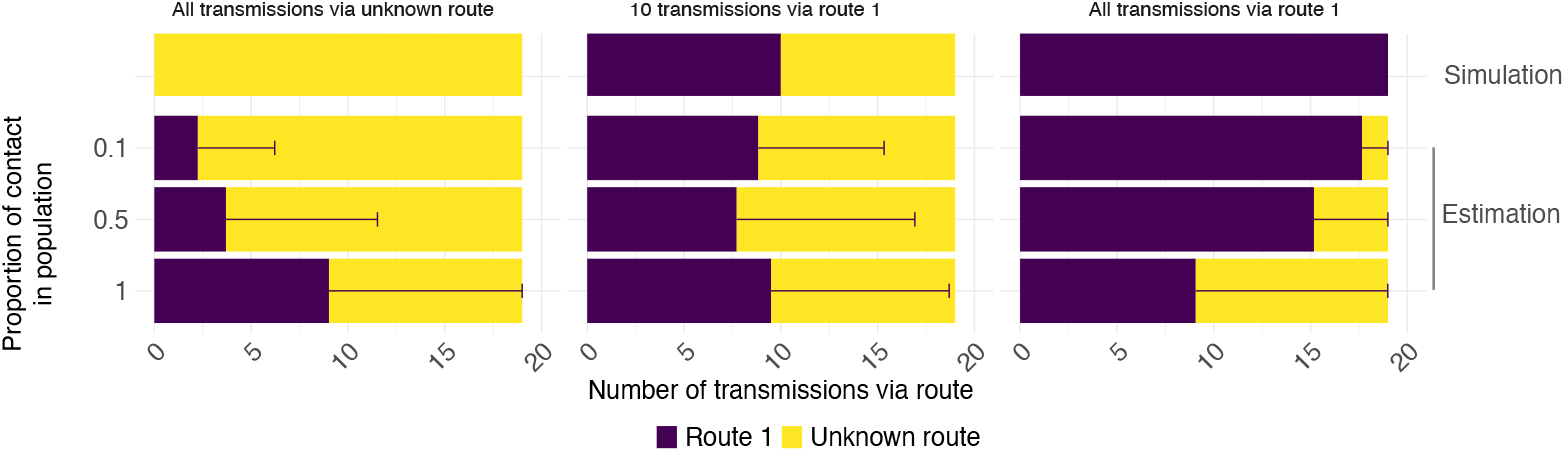
Inference results for 1 known contact type. Estimated number of transmissions via the known transmission route 1 with different proportions of contact of type 1 in the population. Error bars give 95% credible interval of number of transmissions via route 1.

In the second group of scenarios, we used the same 25 simulated outbreaks as before and constructed for each outbreak two contact data matrices for two known transmission routes. The presence of a contact type was independent of the presence of the other contact type. The parameter values to construct the contact data are again found in Table 1. As with one known contact type, the model inferred the simulated number of known transmissions reliably when the contact data was sparse, i.e. proportion of known contact was 0.1 for both types (Fig 3). This was the case when transmissions were simulated to occur via only one route, via two routes and when the unknown route was the most prominent. When the proportions of both types of contacts increased, the estimated numbers of transmissions were again equally divided across all routes, here one third of all transmissions.

**Fig 3.**
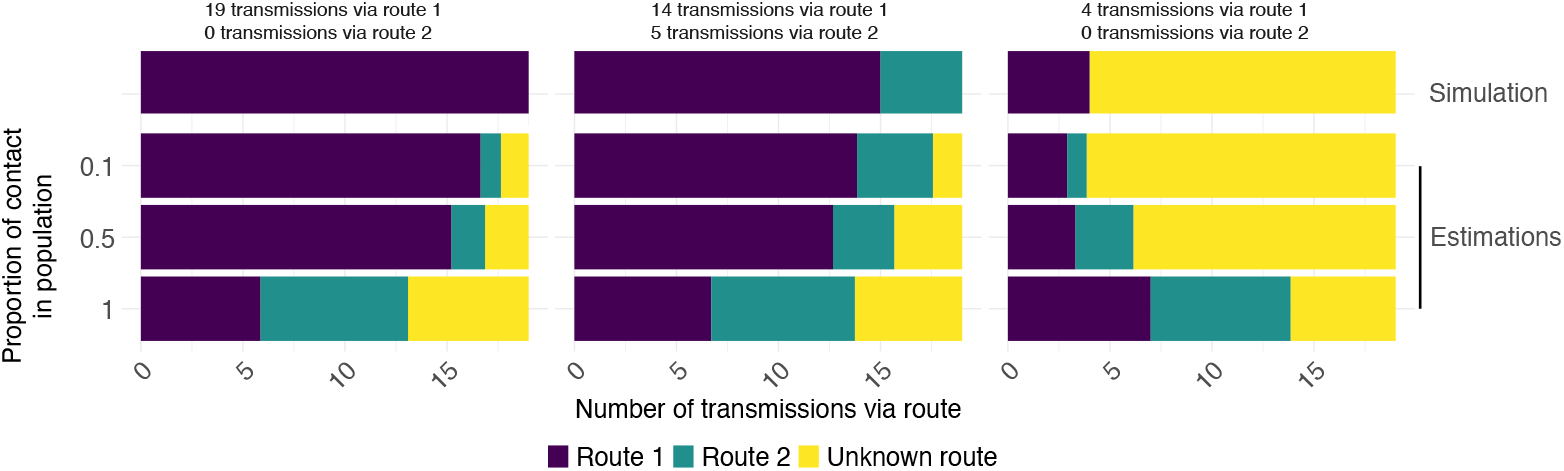
Inference results of number of transmissions via two determined transmission pathways. Both simulated and estimated numbers are shown for different levels of importance of the transmission routes (columns) and proportions of contact (rows). Proportion of contact is the same for both known contact types. The 95% credibility intervals can be found in Table in S1 Table.

The final group of scenarios consisted of two contact types, the presence of which was either perfectly positively correlated or perfectly negatively correlated. Perfect positive correlation means that a contact of type A always coincides with a contact of type B, so there is no information to distinguish the two transmission pathways. Parameter values for the simulation of contact data matrices are again found in Table 1. The model indeed equally divided the total number of transmissions over both known transmission routes (Fig 4). When the unknown route is the most prominent, the remaining transmissions were divided over the two known routes. However, when the two known types were perfectly positively correlated but the proportion of contact type 1 was 5 times higher than the proportion of contact type 2, the data do contain information about the contribution of the two transmission routes to transmission, and the estimated numbers were close to the simulated (Fig 4 -unequal contact proportions). Perfect negative correlation means that no host pair has contacts of both type A and B. The inference results showed that for negatively correlated contact data, the estimated numbers of transmissions via the known routes are close to the simulated numbers. Scenarios with independent occurrence of contacts and those with perfectly negatively correlated contacts showed similar results according to the 95% credible intervals of the number of transmissions via known routes (Table in S2 Table).

**Fig 4.**
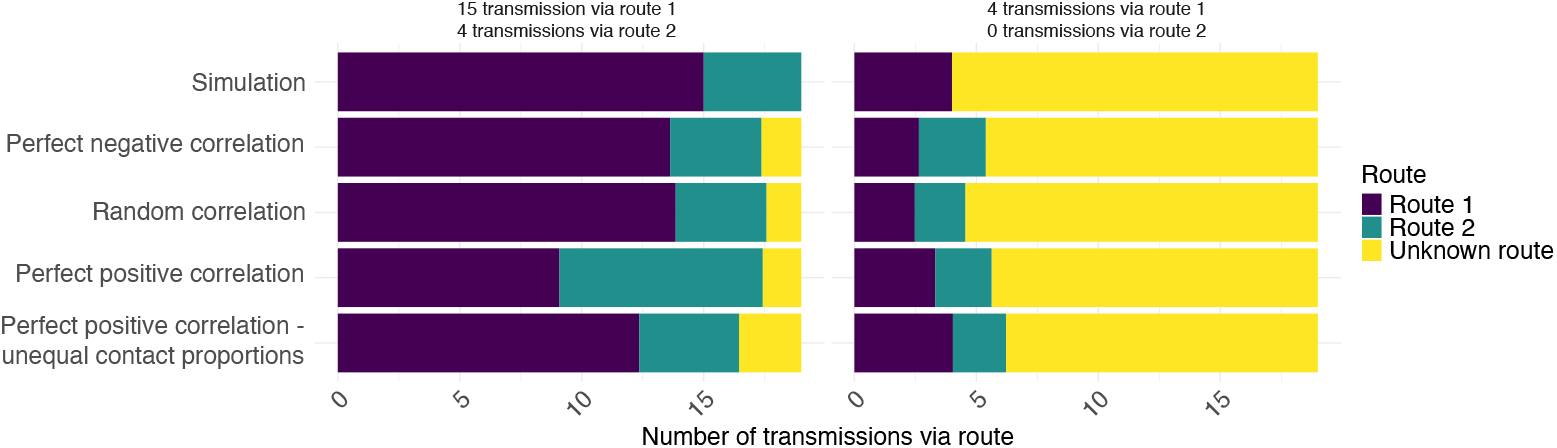
Inference results of numbers of transmissions via two known transmission pathways, where contacts are perfect negatively correlated, random correlated, perfect positively correlated, or perfect positive correlated, with a proportion of contact of route 1 five times less than proportion of route 2. Two scenarios with different numbers of transmissions via the known routes are shown. Proportion of contact is the same for both known transmission routes, except where stated otherwise.

### Enhancing the transmission tree inference by contact data

In addition to the quantification of the importance of contact types, we assessed the fraction of correctly identified infectors from the inference results of the simulations of scenario groups 1 and 2. Based on these fractions, using contact data improved inference when contact was sparse and important for transmission, i.e. large numbers of transmissions occurred via the known route (Fig 5A). This large fraction of correctly identified infectors decreased slightly when the transmissions were split over two known routes, the fraction was higher for 19 transmissions via route 1 than for 15 transmissions via route 1 and 4 via route 2 (Fig 5B).

**Fig 5.**
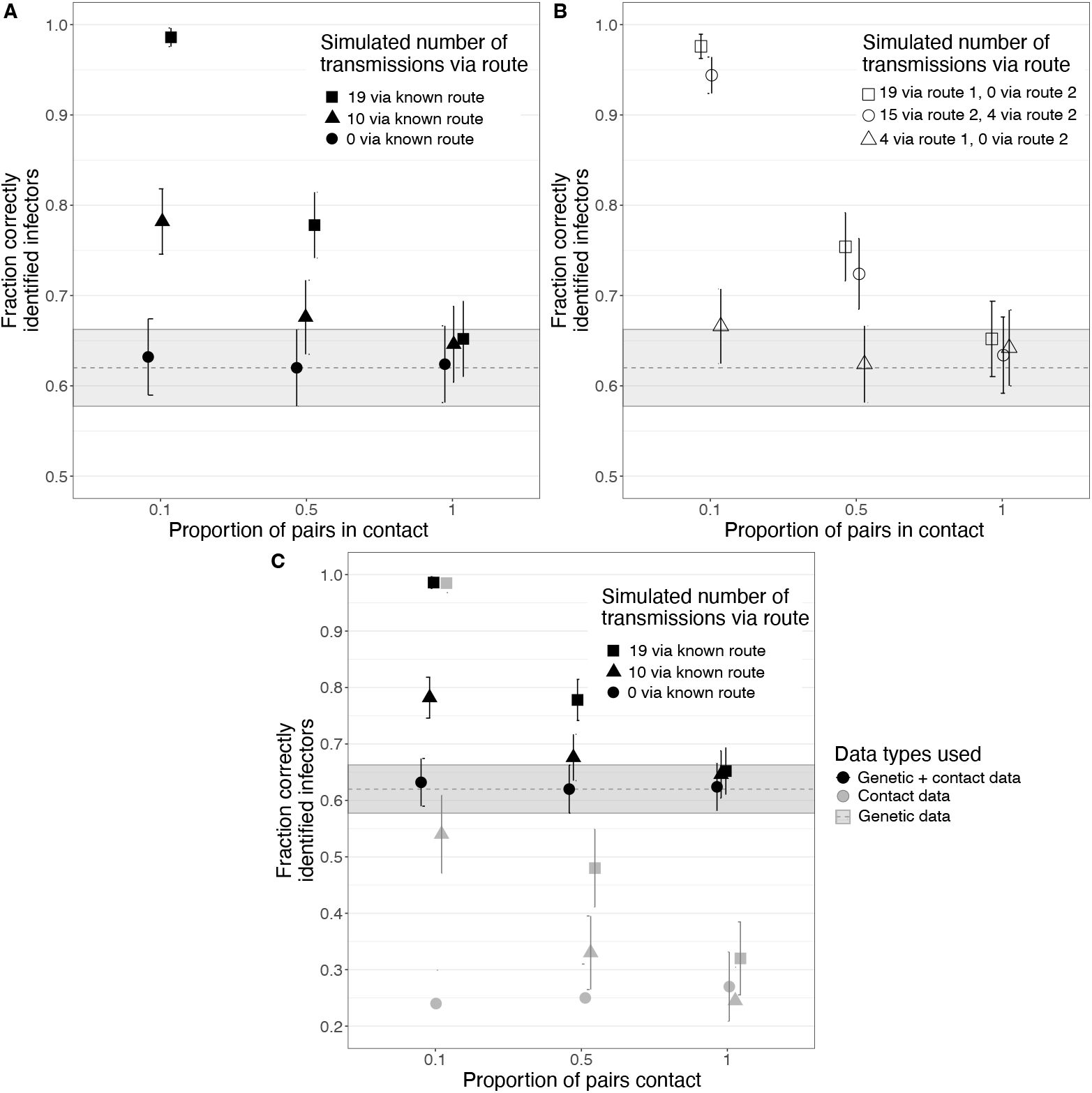
Fractions of correctly identified infectors. Grey dashed lines and shaded areas in all panels represent results of inference without contact data. A) Simulations with 1 known contact type and 0, 10 or 19 transmissions occurring via this type. Proportion of contact in population simulated at 0.1, 0.5 or 1. B) Simulations with 2 known contact types and combinations of transmissions via these types. Proportion of contact in population for both types simulated at 0.1, 0.5 and 1. C) Genetic and contact data used in the inference (black) or only contact data (grey). Simulations with 1 known contact type and 0, 10 or 19 transmissions occurred via this type. Proportion of contact in population simulated at 0.1, 0.5 or 1.

When a contact type is sparse yet important for transmissions, inference with only contact data outperforms inference with only genetic data (Fig 5C). Otherwise, when the contact type is less important and/or not sparse, inference is better when using genetic data alone or in combination with contact data. Another way to enhance the inference is to use informative priors on the importance of contact types. The estimation of number of transmissions via the known transmission route will improve, even when contacts are dense, e.g. the proportion of contact is 1 (Figure in S3 Figure). Nevertheless, the fraction of correctly identified infectors is similar to the fraction found by inference with the uninformative prior.

### SARS-CoV-2 outbreak in Dutch mink farms

During the SARS-CoV-2 pandemic in 2020, 62 of 168 mink farms in the Netherlands were infected. By means of phylogenetics, these farms were divided in 5 genetic clusters [13]. Transmission inference gave evidence to split these genetic clusters in a total of 13 transmission chains [14]. Epidemiological data was collected about the veterinary service provider, the feed supplier, and any personnel link, where farms had either the same owner, they shared employees or are owned by other members of the same family. These data were checked for correlation, i.e. whether two types of contact were observed together more often than expected at random (Table 2). All combinations of contact types occurred more often than expected if they were randomly distributed, suggesting almost perfect positive correlation between all contact types.

**Table 2.**
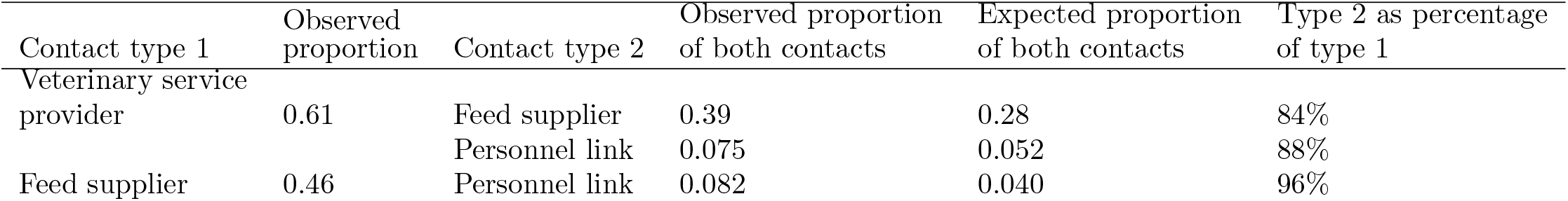
Proportions of farm pairs with multiple types of contact. The expected proportions are computed for contacts distributed at random.

| Contact type 1 | Observed proportion | Contact type 2 | Observed proportion of both contacts | Expected proportion of both contacts | Type 2 as percentage of type 1 |
| --- | --- | --- | --- | --- | --- |
| Veterinary service provider | 0.61 | Feed supplier | 0.39 | 0.28 | 84% |
|  |  | Personnel link | 0.075 | 0.052 | 88% |
| Feed supplier | 0.46 | Personnel link | 0.082 | 0.040 | 96% |

We estimated a mean number of introductions of 11 (IQR: 9-13). The transmission tree corresponding to the maximum parent credibility phylogenetic tree is shown in Figure in S4 Figure. Using the recorded contact types, we estimated the number of farms infected via the corresponding transmission routes. On average 5.1, 5.3 and 6.2 farms were infected via respectively the veterinary service provider, the feed supplier and the personnel link (Table 3). We also computed which fraction of transmission pairs had contact of these types, and in what percentage of these transmission pairs transmission occurred through these contact types. In 76% of all transmission pairs with a personnel link, we expected the farms to be infected via this route, so we conclude that personnel link was a risky contact type for transmission. For the shared veterinary service provider or shared feed supplier, only 19% and 23% of the inferred transmission pairs with the respective types of contact were expected to transmit to each other via these routes.

**Table 3.** Expected number of mink farms infected by SARS-CoV-2 per infection route (fraction of all infected farms). Percentage of observed number of contacts of route for the inferred transmission pairs (expected number of farms where infection occurred via route / number of transmission pairs with this contact type)

|  | Veterinary service provider | Feed supplier | Personnel link | Introduction | Undetermined |
| --- | --- | --- | --- | --- | --- |
| Expected number of farms where infection occurred via route | 5.1 (0.08) | 5.2 (0.08) | 6.2 (0.10) | 11 (0.18) | 34.4 (0.55) |
| Percentage of observed contacts between inferred transmission pairs | 19% (5.1/26.3) | 23% (5.2/23.1) | 76% (6.2/8.2) | - | - |

To quantify the effect of contact data on the certainty of the transmission events, we calculated the entropy of the inference results with and without contact data, which were 71.5 and 74.5 respectively. So the use of contact data slightly increased the certainty of the transmission events.

## Discussion

Understanding the importance of different types of contact for transmission is key to targeting interventions to these contacts to prevent transmission. In this study, we extended the `phybreak` model for transmission tree inference to quantify the contributions of different types of contacts to transmission. Data on contacts of different types between infected hosts are used in addition to genetic sequence data and information on sampling times of infected hosts. Not only is the importance of known contact types quantified, but also their contributions to transmission are compared to the contribution of unknown transmission routes. Using simulated data, inference with the extended `phybreak` model showed that it can reliably estimate the number of transmissions from scarce contact types. We observed this for one (Fig. 2) and multiple known types of contact (Fig. 3). However, if the proportion of hosts connected via known types of contacts is too large, the importance of these contacts cannot be well distinguished from the unknown contact which connects all hosts with each other. In case of perfectly positively correlated contact types - and with equal proportions of contact for all types - all contact types are estimated to be equally important (Fig. 4). Perfect negative correlation results in better estimates of the importance of contact types compared to independent presence of contact. We also found that using contact data as an additional source of information above sequence and other epidemiological data enhanced the transmission tree inference, especially when these contacts were sparse and were important for transmission (Fig. 5).

Applied to data from a SARS-CoV-2 outbreak on Dutch mink farms, we quantified the importance of the following contact types: having a common veterinary service provider, a common feed supplier, or sharing personnel. Transmissions using these contact types accounted for an average of 16.4 of the 62 infections, with transmission occurring via the personnel link for 76% of the infected farm pairs which had a personnel link. This aligned with previous findings showing that personnel links were positively correlated with transmission [13] [15]. For the veterinary service provider and feed supplier we found that on average 26.3 and 23.1 transmission pairs had contact of one of these types. However, only in 5.1 and 6.3 transmission pairs, respectively, transmission occurred via these contacts. So these contact types are found less important using our method than expected based on the inferred transmission pairs and their contacts. Moreover, our simulations showed that when about half of the host pairs had two types of contact, but very few transmissions actually occurred through these contacts, the contribution of one contact type was overestimated by 12.5% (Fig.3). For the mink farms, this would correspond to roughly six transmissions, suggesting that the role in transmission of the veterinary service provider or feed supplier may be overestimated here.

Mink farming is abolished in the Netherlands since this outbreak, but similar questions on which are important transmission routes between infected farms, are relevant for other farm animals and their diseases [16] [17] [18]. Usually data are available about veterinary service providers, feed suppliers, and personnel. For human infectious diseases our methods could be applied to quantify the contribution of different locations to transmission of a pathogen (e.g. hospital wards, meeting locations), co-workers or health care personnel or infected surfaces [19] [20] [21].

A limitation of our method is that we did not model the reporting process of the contact data. Essentially, we estimated the contribution of ‘reported contacts’ to the likelihood of transmission. This means that generalizability of the importance of contact types is dependent on the underlying reporting probabilities. If reporting happens at random for all pairs, i.e. the probability of reporting a contact for a transmission pair is equal to the probability of reporting a contact for a non-transmission pair, then the results are generalizable. However, when the reporting probability of a contact type is higher for transmission pairs, then the importance of that contact type is overestimated and vice versa. Campbell et al. [11] extended the `outbreaker2` inference model to estimate the fraction of reported contacts, allowing for inference on incomplete contact sets. However, implementing that in `phybreak` with multiple types of contact would result in too many parameters to be estimated. Further research may focus on implementation of reporting contacts with multiple contact types.

Possible extensions of the model are to include information about the timing of contacts and the directions of contact. When information about timing of contacts for each contact type is available, e.g. when a veterinarian visited a farm or when a patient was on a hospital ward, the time when transmission could have occurred via that type of contact is restricted to specific time points or specific time periods. This knowledge increases the information on that contact type and can thus refine the estimation of the importance of the contact type. We also assumed that contacts are undirected. Directed contacts - contacts in which the transmission can only go in one direction, e.g. farms that are always serviced in the same order by feed suppliers - may also exclude certain transmission routes and therefore increase the available information about transmission. Again, the importance of contact types may then be refined.

In conclusion, we extended the `phybreak` model to include data on contact between infected hosts in the inference. Using such data, the contribution of different types of contact to transmission can be quantified. In addition, we demonstrated that incorporating contact data provides valuable information to improve transmission tree inference. Our method may help to identify important transmission routes in outbreaks. This knowledge can be used to design interventions that are targeted to high risk contacts and therefore prevent future outbreaks.

## Methods

In this study, we used a Bayesian inference model to infer the phylogenetic tree and transmission tree of an infectious disease outbreak and to quantify the importance of contact types using the corresponding contact data. To do this, we extended the phybreak model [14], such that contact data can be incorporated into the inference as an additional source of information. By contact data we mean information on whether or not pairs of hosts in the data set are connected via contacts of specific types. In other words, contact data are described by matrices with matrix elements being 0 or 1 depending on whether or not a contact between two hosts was observed. For every type of contact, a separate contact matrix is assumed to be available. The model describes four processes on which the transmission tree is inferred: 1) The observation of the pathogen through sampling of genetic sequences, 2) the dynamics of the pathogen that occur within infected hosts and the history host, 3) genetic mutations in the pathogen and 4) infection of cases from outside the population or via transmission from other cases. The process of transmission is discussed here. All other processes are discussed in the Supplementary materials.

### Transmission pathways affects the transmission process

The transmission process is described by a Poisson process. We assume that infected cases are generated in an infinite susceptible population. The incidence of cases subsequent to the initial introduction of a pathogen is modelled by two distinct processes: additional introductions from outside the population of infected hosts and transmission between hosts. Additional introductions occur with exponentially distributed waiting times, characterized by a rate *λ*_intro_, after the initial introduction until the final sample time. The time between the first introduction and the last sample time is denoted by *T* and the number of introductions by *k*. The transmission process is characterized by a dynamic rate, depending on the times since infection of infected hosts, as described by the generation time distribution. This distribution is modelled as a Gamma distribution with shape parameter *a*_*G*_ and mean *m*_*G*_. The probability density function of the generation time of a host *i*, with 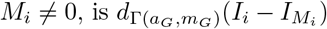 where *I*_*i*_ and *I*_*M*_*i* are the infection times of respectively host *i* and its infector host *M*_*i*_. The reproduction number, i.e. the average number of secondary cases per primary case, is assumed constant during the outbreak and denoted by *R*. The likelihood for the transmission tree is:

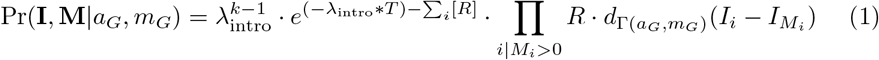

Assume that we know all contacts of *n* different types, we then also have *n* known transmission routes. For each contact type *r*, the probability of pairs of hosts to be connected by this type is denoted by *p*^*r*^. The contacts are stored in a binary symmetric matrix *A*^*r*^ and the expected fraction of transmissions that have occurred via a route *r* is denoted by *x*_*r*_. As transmission might have happened via contact types not in the data, we also include an unknown transmission route *r* = 0. For this route, we assume that each host is connected with each other host, such that *p*^0^ = 1 and *A*^0^ contains only 1’s.

Now assume the following: 1) The proportion of contacts between non-transmission pairs, i.e. pairs of infected hosts that did not transmit the pathogen to each other, is equal to *p*^*r*^. 2) The probability of a contact of type *r* between any two hosts is independent of host characteristics, but may depend on presence of a contact of type *s* ≠ *r*, e.g. farms with the same owner may also have the same feed supplier. 3) Contacts do not change over time, such that *A*^*r*^ = *A*^*r*^(*τ* ), ∀*τ* ∈ [0, *T* ]. We can write the reproduction number *R* as:

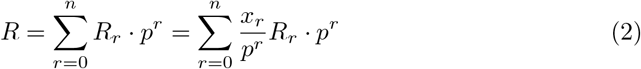

where *R* is expressed as the sum of the contributions by all contact types, which are then expressed as the product of the relative transmissibility of each contact type (*x*_*r*_*/p*^*r*^) and the frequency of the contact type in the population *p*^*r*^. We can use the relative transmissibilities of the contact types to weigh transmission events between hosts *i* and *M*_*i*_ in the likelihood (Eq 1), and obtain:

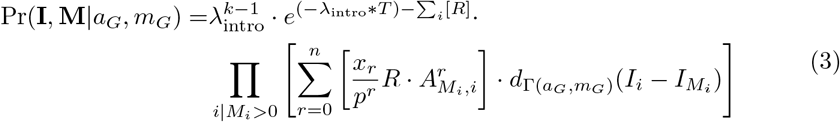

In the product term we look at the infection pressure of host *M*_*i*_ on host *i* which is the sum of contributions *R*_*r*_ over all contacts *r* that were present between *i* and *M*_*i*_ .

The MCMC chain let us now obtains samples from the posterior distributions of *x*_*r*_, i.e. the expected fraction of transmissions via route *r*. The prior expected fractions of transmissions *x*_*r*_ has a Dirichlet distribution, with *α*_*r*_ = 0.5 for all contact types *r* (‘uninformative’ Jeffrey’s prior), or *α*_*r*_ = 10 * (1 −*x*_*r*_) (informative prior). With only one contact type in the data, these are equivalent to Beta distributions.

To reduce the number of parameters in the MCMC we chose to compute the proportions of contact *p*^*r*^ directly from the contact data matrices by

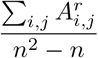

where *n* is the number of hosts. In this way, contacts of transmission pairs are contributing to the total proportion of contact *p*^*r*^. As we assumed that the population-wide proportion of contact is equal to the proportion of contact in the infected non-transmission pairs, we overestimate *p*^*r*^ with a maximum of 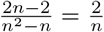 when all transmission pairs have contact of type *r*.

### Simulation with transmission pathways

To assess the performance of the model with multiple transmission pathways, we simulated outbreaks and constructed for each route *r* ≥ 1 a corresponding contact data matrix *A*^*r*^ given the population-wide proportion of contact *p*^*r*^ and the expected fraction of transmissions via this route *x*_*r*_. Assume that the transmission route for a infector-infectee pair (*M*_*i*_, *i*) is *r*. Then 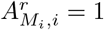, because contact has to be present for transmission. Since transmission between *M*_*i*_ and *i* is now no longer possible via another route *s* ≠ *r*, occurrence of contacts of type *s* ≠ *r* between *M*_*i*_ and *i* are sampled with probability *p*^*s*^. As we sample a given number of transmissions for each route *r*, the order in which types of contacts are allocated does not matter. All non-transmission pairs (*j, i*) have a contact of type *r* with probability *p*^*r*^.

We simulated three groups of scenarios: 1) one known contact type *r* = 1; 2) two known contact types *r* = 1, 2 in which presence of contact of one type is independent of presence of the contact of the other type; 3) Two known contact types *r* = 1, 2 with *p*^1^ ≥ *p*^2^, in which presence of contact type 2 always leads to presence of contact type 1 (perfect positive correlation) or presence of contact type 2 always leads to absence of contact type 1 (perfect negative correlation).

In total, 25 outbreaks were simulated with 20 hosts and 1 index case, such that there are 19 transmissions in total. Simulation of an outbreak was done by creating a transmission tree via a Poisson model where the generation time between two hosts is Gamma distributed with a shape of 10 and mean of 1 [14]. After infection, the sampling of a host takes place after a Gamma distributed time with a shape of 10 and a mean of 1. A phylogenetic tree is constructed on the transmission tree using a coalescence rate of 1. For the genetic sequences, we sampled the number of mutations from a Poisson distribution with parameter equal to *λ* = *μ* · sequence length · total length of all edges, where *μ* = 1 · 10^*−*4^ is the mutation rate per time unit and the sequence length is 10^4^. The mutations are distributed over the edges of the phylogenetic tree, with the lengths of the edges as weigths. On average, 1 mutation per genome per time unit occurred.

During the inference, the number of introductions was inferred with the model by obtaining the most likely transmission tree. For all scenarios, the same 25 outbreaks were used on which to construct the contact data matrices. The values of *p* and *x* used in the different scenarios to construct the contact data are given in Table 1, where for *x* we give the number of transmissions via a transmission route (19 times the fraction of transmissions).

To assess the performance of the method, we compared the estimated numbers of transmissions that occurred via pathways, with the simulated numbers of transmissions. In addition, we calculated the accuracy, which is the fraction of correctly identified infectors from the inference.

### Real world example: SARS-Cov-2 in Dutch mink farms

We previously described an outbreak of the SARS-CoV-2 virus in the Dutch mink farms in the Netherlands in 2020 [14]. We obtained 326 whole-genome-sequences of SARS-CoV-2 sampled in minks from the GISAID database, along with their time of sampling. After alignment and removal of positions with only N, all sequences were 16,289 nucleotides long. Metadata of the farms were collected by the One Health mink outbreak investigation consortium and included the time of culling of the farms, the veterinary service provider and the feed company of the farm [13]. Farms were also grouped by sharing either the same owner, same personnel or any family link. We created 3 contact matrices were two farms are connected by contact if they have 1) the same veterinary service provider, 2) the same feed company or 3) a personnel link, i.e. farms sharing personnel, having the same owner, or with the owners being family members. The contact data matrices were checked for correlation. The proportion of host pairs which had two contact types was compared to the expected proportion with two contact types, calculated by multiplying the proportions of contact in the two separate contact matrices.

As in a previous analysis of the same outbreak [14], we assumed that infectiousness of the mink farms follows a logistic curve, with exponential decay after culling at time *T*_*c*_:

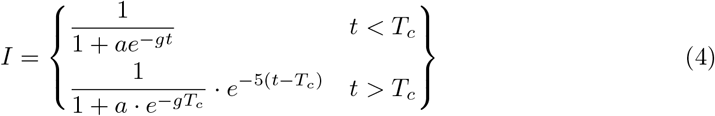

Here, *a* = 1 · 10^*−*4^ is the initial part of the mink population at a farm being infected, *g* is the growth rate, and *t* is the time after infection of the farm. The logic behind the exponential decay after culling is to have a delay in clearing of infectiousness, for instance due to environmental contamination. Because the value for *T*_*c*_ differ per farm, the infectiousness curves differ between the farms. Therefore we normalize the curves, such that the average infectiousness function integrates to 1, while accounting for higher total infectivity of longer infected farms.

Two MCMC chains with 20,000 steps were run for an Effective Sample Size (ESS) above 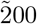 of all model parameters to ensure convergence. Furthermore, a MCMC chain of length 20,000 steps was run without contact data to assess the effect of the contact data on the inference. Entropy in the set of infectors of the farms was calculated as a measure of uncertainty about the infector, using the following equation [22]:

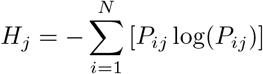

with *P*_*ij*_ the support for farm *j* to be infected by farm *i*, with *N* possible infectors. If farm *j* has one infector with support 1, the entropy will be 0. Using the entropy, we compared the inference results with and without contact data to see if inclusion of contact data provided more certainty about the transmission tree (lower entropy).

## Data Availability

The whole-genome-sequences of SARS-CoV-2 sampled in minks were obtained from the GISAID database. Metadata of the farms is published by Lu et al. (2021, doi:10.1038/s41467-021-27096-9).

## Acknowledgments

We gratefully acknowledge funding by the The Netherlands Organisation for Health Research and Development ZonMw grant number 10710022210003. This work was also supported by the Netherlands Center of One Health (www.ncoh.nl).

